# Imaging Blood-Brain Barrier Dysfunction in Schizophrenia Spectrum Disorders: protocol of the longitudinal IMPACT DCE-MRI study

**DOI:** 10.64898/2026.01.09.26343751

**Authors:** Anne Wendl, Amelie Zöllinger, Daniel Lukas, Isabel Lutz, Michelle Schamberger, Constanze Lobinger, Isabel Maurus, Alkomiet Hasan, Elias Wagner, Peter Falkai, Andrea Schmitt, Florian J. Raabe, Kolja Schiltz, Emanuel Boudriot, Vladislav Yakimov, Lukas Roell, Daniel Keeser, Joanna Moussiopoulou

## Abstract

Schizophrenia spectrum disorders (SSDs) are clinically and biologically heterogeneous and lack reliable biomarkers for stratification and outcome prediction. Evidence from postmortem, fluid biomarker, and neuroimaging studies suggests that blood–brain barrier (BBB) dysfunction may contribute to pathophysiology in a biologically defined subgroup. However, findings are inconsistent and often based on cross-sectional or indirect measures.

The IMPACT study is a longitudinal, multimodal investigation designed to quantify BBB permeability across disease phases using dynamic contrast-enhanced magnetic resonance imaging (DCE-MRI) and to integrate these measures with deep clinical phenotyping. We recruit inpatients with SSDs and healthy controls (HC). Participants undergo multimodal MRI including DCE-MRI at three time points: acute psychosis (baseline; V1), early treatment (4-6 weeks; V2), and long-term follow-up (2.5 years; V3). Clinical characterization includes standardized measures of psychopathology, functioning and cognition. Blood samples are collected at each visit, while CSF is obtained only when clinically indicated in the SSD group. DCE-MRI-derived voxelwise permeability metrics are then analyzed.

Primary objectives are to (i) compare BBB leakage cross-sectionally between SSD and HC and (ii) characterize spatial and temporal leakage profiles across illness phases. Secondary objectives include analyzing associations with psychopathology and biological (e.g., inflammatory) signatures, as well as exploratory identification of subgroups. By providing a longitudinal, BBB-specific neuroimaging framework embedded in a deep phenotyping infrastructure, IMPACT aims to elucidate BBB alterations in SSDs and to support biologically informed stratification approaches in precision psychiatry.

## 1. Introduction

Patients with schizophrenia-spectrum disorders (SSDs) show heterogeneity in symptom profiles, development of functional capacity and responsiveness to treatment, ranging from isolated acute episodes to chronic, treatment-resistant illness [1]. Despite decades of research into neuroimaging, fluid biomarkers and genetics, reliable biological markers for diagnosis or treatment guidance are still lacking and the underlying pathophysiology remains incompletely understood [2]. Amongst emerging biological systems of interest, the blood-brain barrier (BBB) gains greater recognition [3]. As a highly selective interface formed by endothelial cells, pericytes and astrocytic end-feet, the BBB strictly regulates molecular and cellular exchange between the systemic circulation and the central nervous system (CNS) [4]. Dysfunction of this barrier can permit entry of peripheral inflammatory mediators or immune cells into brain tissue, potentially initiating neuroinflammatory and degenerative processes implicated in neuropsychiatric disorders [5]. Although BBB deficiency is documented in neurological disorders such as Alzheimer’s disease, Parkinson’s disease, multiple sclerosis [6], its role in SSDs remains only partially elucidated.

Evidence from postmortem analyses and in vivo assessments of blood and cerebrospinal fluid (CSF) parameters have been postulated to indicate alterations of the BBB in SSDs. Postmortem investigations reveal structural and morphological changes within the neurovascular unit [7], including reduced numbers of pericapillary oligodendrocytes [8], endothelial vacuolation, thickening of the basal membrane, and hypertrophy of astrocytic end-feet [9]. However, these data are restricted by small sample sizes, potential biases related to the cause of death, and the inability to capture in vivo or longitudinal dynamics in BBB changes. Fluid-based biomarkers such as the CSF to serum albumin ratio (Q_Alb_), widely regarded as state-of-the-art method for assessing BBB dysfunction [3], have been reported elevated in individuals with SSDs [10, 11]. However, this marker predominantly reflects the function of the blood-CSF barrier (BCSFB) rather than the BBB, and is susceptible to extracerebral influences and other confounding factors [12].

Dynamic contrast-enhanced MRI (DCE-MRI) is a highly sensitive technique for quantifying even subtle BBB leakage and has demonstrated utility across various neuropsychiatric conditions, including Alzheimer’s disease [13, 14]. BBB disruption can be measured from the extravasation of intravenously administered contrast agent (CA), which results in image contrast enhancement [15]. Temporal changes in T1-weighted signal intensity are analyzed using pharmacokinetic models, to derive quantitative permeability metrics [16, 17]. The most widely applied parameter, K_trans_ reflects the rate of CA transfer from plasma to the extravascular, extracellular space [15, 16]. One cross-sectional DCE-MRI study that has assessed BBB permeability in vivo in SSD patients reported increases in K_trans_ values within the thalamus in a cohort of 29 patients compared with 18 healthy controls, but its statistical power was limited due to the small sample size [18]. Consequently, this finding requires replication and more detailed characterization of the affected phenotype. Moreover, the cross-sectional design precludes insight into the temporal dynamics of BBB alterations across stages of illness [3, 18]. A more comprehensive, systematic approach is therefore needed to investigate BBB integrity in SSD and to relate it to clinical phenotype, illness stage, and biologically informed profiles, including peripheral and cerebrospinal fluid inflammatory markers [3, 19]. Such efforts are essential to establish the clinical relevance of BBB dysfunction, identify affected patient subgroups, and evaluate its potential as a target for precision psychiatry and individualized therapeutic interventions [3, 20].

The *Imaging-Blood-Brain-Barrier-Permeability-in-Schizophrenia-Spectrum-Disorders-Study* (IMPACT Study) is a longitudinal, multimodal, naturalistic investigation of BBB function in SSDs,. DCE-MRI is performed at multiple illness and treatment stages to assess spatial and temporal patterns of BBB permeability [13, 16], analyzed via pharmacokinetic modeling [17, 21]. Combined with clinical, cognitive, cerebrospinal fluid and blood biomarkers, as well as additional neuroimaging modalities, this design enables comprehensive biological phenotyping. This study aims to compare BBB leakage between individuals with SSDs and healthy controls; characterize its spatial and temporal profile; examine clinical correlates, including symptoms, cognition, and functioning; investigate underlying inflammatory mechanisms; and evaluate BBB dysfunction as a biomarker for SSD subgroups. Ultimately, IMPACT seeks to advance biomarker-guided, mechanistically informed precision psychiatry [3].

## 2. Methods and design

### 2.1. Study design and setting

IMPACT is a prospective, naturalistic, longitudinal observational study conducted at the Department of Psychiatry and Psychotherapy, LMU University Hospital Munich, with harmonized procedures embedded in the *Clinical Deep Phenotyping of Treatment Response in Schizophrenia (CDP-STAR)* infrastructure [22]. CDP-STAR was approved by the local ethics committee (project number 24-0341, 2024) and is registered at the German Clinical Trials Register (DRKS00034820) and OSF (https://doi.org/10.17605/OSF.IO/SQ2TZ). The IMPACT protocol was independently approved by the local ethics committee of LMU Munich (reference number 21-1139) and is conducted in accordance with the Declaration of Helsinki.

### 2.2. Study population

#### Schizophrenia spectrum disorder group

The study includes inpatients diagnosed with schizophrenia spectrum disorders (SSDs), including both first-episode psychosis and recurrent psychotic episodes, according to the criteria of the Diagnostic and Statistical Manual of Mental Disorders (DSM-5-TR, Version 7.0.2). All diagnoses are confirmed using the Mini-International Neuropsychiatric Interview (M.I.N.I.) [23]. Additional eligibility criteria include an age range of 18 to 65 years, sufficient German language proficiency, the capacity to give written informed consent and inclusion in the Munich Mental Health Biobank (MMHB) [24]. Exclusion criteria comprise a primary psychiatric diagnosis other than SSDs, the presence of acute risk to self or others (e.g., acute suicidality), ongoing coercive treatment or involuntary hospitalization at the time of study inclusion. Participants are also excluded if they are unable to provide informed consent, are currently pregnant, or have relevant comorbid disorders affecting the central nervous or immune system - such as epilepsy, dementia, multiple sclerosis, organic psychotic or affective disorders, or acute inflammatory conditions. Further reasons for exclusion are general contraindications for cranial MRI, such as implanted pacemakers, as well as specific contraindications to dynamic contrast-enhanced MRI (DCE-MRI), including acute renal failure or known hypersensitivity to Gadolinium-based contrast agents. Potential participants are routinely screened for eligibility by trained study physicians at our clinic, and written informed consent is obtained before the initiation of any study-related procedures.

#### Healthy control group

Healthy controls (HC) are recruited and screened to exclude current or past psychiatric disorders (self-report), relevant neurological disease, MRI contraindications, and contraindications to gadolinium administration. Additional exclusion criteria include current use of psychotropic medication. The HC group participates only in a single study visit.

### 2.3. Study timeline

The study protocol is outlined in **Figure 1**. Healthy controls participate in a single study visit, whereas individuals with SSD are invited to complete all three study visits. At baseline (visit 1; V1), all participants undergo a standardized assessment battery comprising multimodal MRI - including DCE-MRI - comprehensive clinical characterization, and venous blood sampling. Cerebrospinal fluid (CSF) is additionally collected via lumbar puncture in the SSD group when clinically indicated, in accordance with German guidelines [25]. Follow-up assessments of the SSD group are scheduled at 4-6 weeks (V2) and 2.5 years (V3) following baseline. These follow-up visits include multimodal and DCE-MRI, clinical and cognitive evaluations, and blood sampling to assess longitudinal changes across the acute phase, early treatment response, and long-term progression of illness.

**Figure 1:**
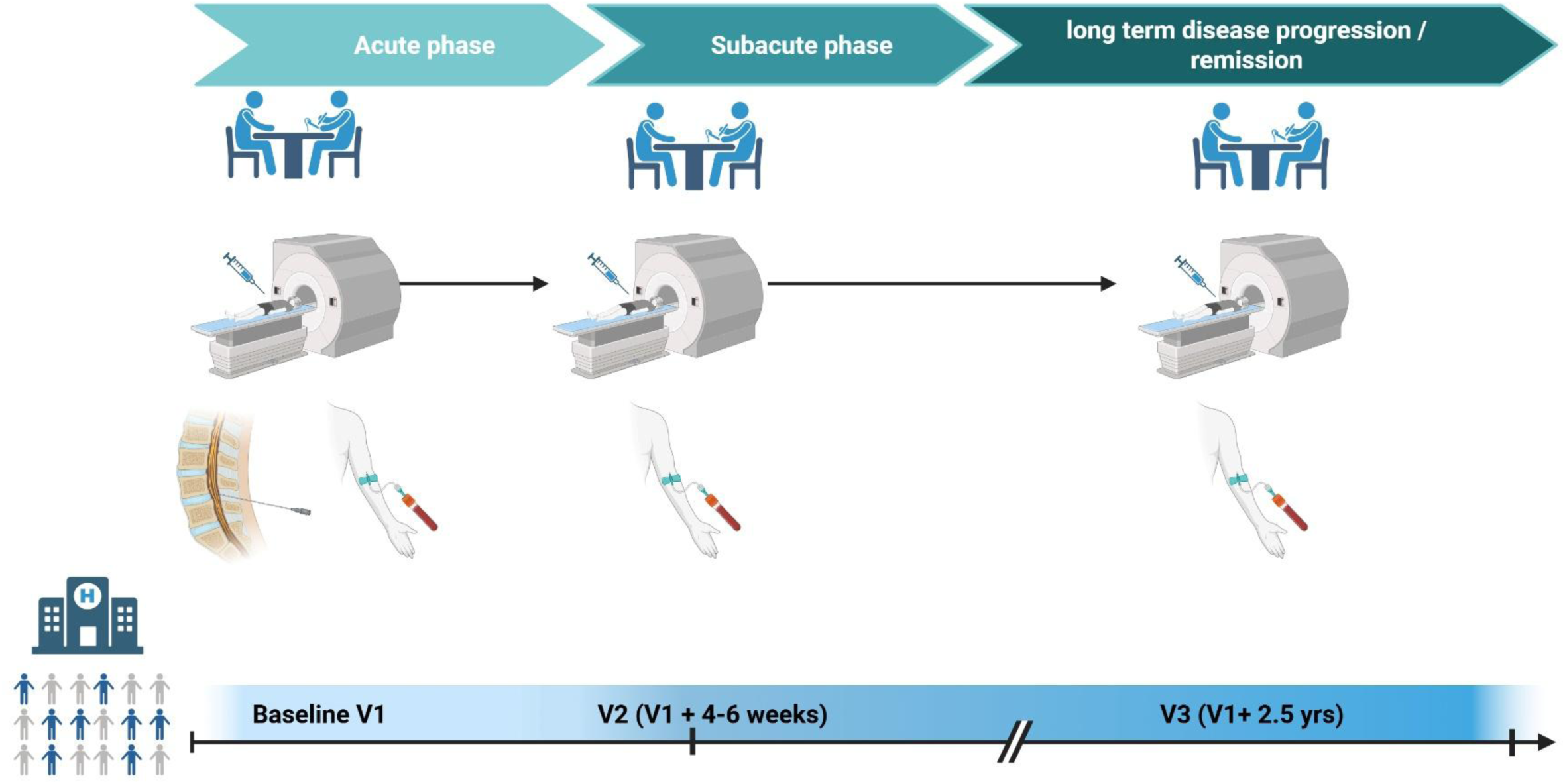
Study design and timeline.

### 2.4. Clinical Characterization

All participants (SSD and HC) complete structured interviews and standardized questionnaires to obtain comprehensive demographic and clinical information. This includes age, sex, height and weight (used to calculate body mass index, BMI), educational background and substance use. Whenever possible, self-reports are corroborated with available medical records to enhance data validity.

The clinical protocol also captures past and present somatic illnesses, with particular attention to neurological and inflammatory disorders and cardiometabolic risk factors, including smoking status, dyslipidemia, and obesity, which are known to contribute to the environmental risk burden in schizophrenia [26].

Participants in the SSD group additionally provide information on current pharmacological treatment and history of electroconvulsive therapy. Further illness-related variables include lifetime exposure to antipsychotic medication (quantified in months, chlorpromazine equivalents), duration of illness (defined as time since first psychiatric diagnosis), age at symptom onset, duration of untreated psychosis, number of psychiatric hospitalizations, and illness stage (first-episode vs. multi-episode psychosis).

Diagnostic validation and psychopathological characterization in individuals with SSD are conducted at the baseline assessment using the Mini-International Neuropsychiatric Interview (M.I.N.I.) based on DSM-5 criteria, which is a validated and efficient tool for structured psychiatric diagnosis in clinical and research settings [23]. Symptom severity and global clinical status are assessed at all three visits using the Positive and Negative Syndrome Scale (PANSS) [27], the Calgary Depression Scale for Schizophrenia (CDSS) [28], the Clinical Global Impression scale (CGI) [29] and the Global Assessment of Functioning (GAF) scale [30]. In addition, participants complete the abbreviated version of the World Health Organization Quality of Life Instrument (WHOQOL-BREF) [31] as part of the V1 and V2 to assess health-related quality of life.

### 2.5. Cognitive Testing

Cognitive performance is assessed in both the SSD and control groups using a standardized test battery administered by trained mental health professionals. To minimize practice effects, different test components are implemented at each timepoint (V1, V2, V3). At baseline (V1), the full battery — comprising the Brief Assessment of Cognition in Schizophrenia (BACS), the Trail making Test (TMT), and Montreal Cognitive Assessment (MoCA) — is administered. At V2 (4-6 weeks), cognitive assessment is limited to the TMT to reduce retest effects, while at V3 both MoCA and TMT are repeated.

The TMT evaluates multiple cognitive domains commonly affected in individuals with SSDs, including attention, visual search and scanning, processing speed, task switching, cognitive flexibility, and executive function [32]. The BACS [33], a validated instrument developed specifically for patients with psychotic disorders, assesses six cognitive domains: verbal memory, working memory, motor speed, attention, executive function, and verbal fluency. These domains align with those identified as central by the Measurement and Treatment Research to Improve Cognition in Schizophrenia (MATRICS) Neurocognition Committee for evaluating cognitive deficits in schizophrenia [34]. In addition, the MoCA is employed to provide a broader evaluation of global cognitive functioning [35]. It assesses a wide range of cognitive abilities, including attention and concentration, executive functions, memory, language, visuospatial abilities, conceptual reasoning, calculations, and orientation.

### 2.6. Multimodal brain imaging

All MRI data are acquired at the Department of Psychiatry and Psychotherapy, University Hospital of LMU Munich, using a 3-Tesla Magnetom Prisma scanner (Siemens Healthcare GmbH, Erlangen, Germany) equipped with a 64-channel head coil. Multimodal MRI (mMRI) acquisition follows the protocol of the Human Connectome Project (HCP) [36], adapted for use in psychiatric populations. Anatomical imaging includes high-resolution T1-weighted magnetization-prepared rapid acquisition gradient echo (T1-MPRAGE) and T2-weighted sampling perfection with application-optimized contrasts using different flip angle evolution (T2-SPACE). Functional imaging includes resting-state functional MRI (fMRI) using a gradient-echo echo-planar imaging sequence in anterior-posterior orientation as well as arterial spin labeling (ASL) for quantitative assessment of cerebral perfusion without the use of contrast agents. DCE-MRI, which forms an integral part of the multimodal MRI protocol, is described in detail in a separate section.

An experienced neuroradiologist reviews all MRI datasets. In cases where structural abnormalities are identified, participants are informed accordingly and receive appropriate clinical care and their scans are excluded from further analysis.

#### 2.6.1. Dynamic contrast-enhanced magnetic resonance imaging (DCE-MRI)

DCE-MRI is included in the multimodal imaging protocol to assess BBB integrity across multiple brain regions. The technique allows quantification of BBB permeability by measuring contrast agent efflux from the vasculature into brain tissue, which is then used to calculate whole-brain voxelwise leakage. The sequence has a total acquisition time of 23 minutes and a voxel size of 1.8 × 1.8 × 1.8 mm. A gadolinium-based contrast agent (Gadobutrol, Gadovist®, Bayer AG, Leverkusen, Germany) is administered intravenously via the antecubital vein at a dose of 0.1 mmol/kg body weight. Injection occurs at time point 12/80 of the scan, approximately 2 minutes after sequence onset, at a rate of 3 ml/s, followed by a 25–30 ml saline flush.

### 2.7. Biobanking of blood and cerebrospinal fluid

All participants need to provide written informed consent for inclusion in the Munich Mental Health Biobank, which provides the infrastructure for our sample biobanking as previously described [3]. All individuals with SSDs undergo basic blood tests including complete blood count analyses as part of the clinical routine in our clinic (collected around 8 a.m.). At baseline (visit 1) the Psychiatry Biobank Blood Kit is collected, containing the following components: 1 × S-Monovette® RNA exact (Sarstedt, Cat. No. 01.2048.001) for RNA extraction, 1 × 7.5 ml K3EDTA tube (Sarstedt, Cat. No. 01.1605.001) for DNA extraction, 1 × 9 ml K3EDTA tube (Sarstedt, Cat. No. 02.1066.001) for plasma-based analysis and 1 × 9 ml tube with coagulation activator (Sarstedt, Cat. No. 02.1063.001) for serum-based analysis. During the DCE-MRI scans one additional 7,5 ml serum tube is collected via the peripheral venous catheter used for contrast agent administration.

As part of standard SSD diagnostics in Germany, CSF examinations are offered to individuals with SSD, in accordance with the national schizophrenia guidelines [37]. Q_Alb_ (as a continuous variable) was selected as indicative for CSF pathology or BCSFB disruption and adjusted for age according to the formula: Q_Alb_ = (4+age/15) × 10−3 [38], age indicated as years. In all cases, CSF and serum are tested for neuronal autoantibodies to rule out autoimmune-mediated psychosis or encephalitis. Long-term storage is performed at −80 °C.

### 2.8. Study progress and power analysis

An a priori power analysis was conducted to estimate the required sample size for the primary cross-sectional comparison of BBB permeability between individuals with SSD and HCs at baseline (V1). As prior DCE-MRI studies in schizophrenia report significant group differences in thalamic K^trans^ but do not provide standardized effect sizes, a moderate effect size was predefined for planning purposes (Cohen’s d = 0.47). This assumption was informed by DCE-MRI studies in ageing, mild cognitive impairment, and Alzheimer’s disease reporting effect sizes in the range of d ≈ 0.4–0.7 for BBB leakage metrics [39], as well as by reports of BCSFB dysfunction based on age-adjusted Q_Alb_ in SSDs [40].Using a two-tailed independent-samples t-test with α = 0.05 and power (1–β) = 0.80, the required sample size was estimated at minimum n = 57 per group (total N = 114). Recruitment commenced in March 2023. As of January 2026, N = 110 individuals with SSD and N = 54 HC (total N= 164) have completed the baseline assessment (V1), thereby meeting the predefined requirements for the primary cross-sectional analysis and allowing additional power for within-SSD analyses. Results addressing the primary objective of baseline BBB permeability differences between SSD and HC have been published elsewhere and confirmed significantly increased BBB leakage in SSDs [41].

Following confirmation of baseline group differences, the study protocol was expanded to include the longitudinal follow-up assessments at 4–6 weeks after baseline (V2; early treatment phase) and at 2.5 years (V3; long-term outcome phase). Longitudinal analyses focus on within-subject changes in BBB permeability and are conducted using linear mixed-effects models.

Because no prior data are available on intra-individual correlations of BBB permeability metrics based on DCE-MRI across time in SSDs, longitudinal analyses are designed as a proof-of-concept. A target sample size of approximately N ≈ 30 SSD participants completing V1 and each follow-up visit (V2 and V3) was defined to allow detection of small-to-moderate within-subject effects (Cohen’s d ≈ 0.4–0.5) at α = 0.05 in repeated-measures designs, while remaining feasible within a naturalistic clinical setting.

Expected attrition rates of approximately 15% at V2 and 30% at V3 were incorporated into the sampling strategy and closely match observed dropout rates. As V2 was implemented later (May 2025), an additional recruitment target of N = 35 SSD participants was defined to ensure approximately N ≈ 30 completers at V2, resulting in a revised target of N = 128 individuals completing V1. As of January 2026, N = 10 SSD participants have completed V2, and N = 20 have completed V3.

All power assumptions and sampling targets were cross-validated against prior longitudinal neuroimaging studies conducted at our site and reflect a balance between statistical rigor, feasibility, and the constraints of a deeply phenotyped, naturalistic cohort.

## 3. Discussion

Psychiatry continues to face challenges in classifying mental diseases beyond symptom-based criteria, lagging behind fields such as oncology or rheumatology, where biomarker-guided treatment is already established [42]. The lack of validated biological markers remains a major barrier to implementing precision medicine in psychiatry [43]. Emerging evidence suggests that blood–brain barrier (BBB) dysfunction may represent a relevant biological mechanism in schizophrenia spectrum disorders (SSDs) and contribute to clinical heterogeneity, treatment response, and illness trajectory [44]. However, existing evidence is limited by indirect measures, small sample sizes, and predominantly cross-sectional designs.

The IMPACT study addresses these limitations by implementing a longitudinal DCE-MRI framework embedded in a deep phenotyping infrastructure. Prior work from our group demonstrates that integrating multimodal data can yield biological insights into SSD pathophysiology and establish associations with clinically relevant outcomes [45, 46]. The IMPACT protocol integrates quantitative BBB permeability mapping with standardized assessments of psychopathology, functioning, and cognition, as well as repeated biosampling (blood at all visits; CSF when clinically indicated). This facilitates the characterization of BBB leakage at clinically meaningful stages (acute psychosis, early treatment, long-term follow-up) and the investigation of cross-sectional and longitudinal associations with clinical and biological profiles. Crucially, the design facilitates analyses beyond case–control comparisons, allowing for the investigation of within-SSD heterogeneity and distinct subgroups. These methodological strengths position the study to support the evaluation of BBB-based stratification in SSDs.

Initial cross-sectional analyses [41] demonstrated increased BBB permeability in SSD compared with healthy controls, supporting feasibility and motivating the longitudinal extension. The longitudinal component is designed to determine whether BBB permeability exhibits state-related dynamics associated with symptom change and treatment, or whether it reflects a stable trait-like characteristic. Furthermore, the longitudinal structure allows future evaluation of whether baseline BBB permeability predicts subsequent clinical and functional outcomes, acknowledging that these analyses are exploratory at the current stage.

Key methodological strengths of the study include: (i) repeated DCE-MRI within the same individuals across defined illness phases; (ii) harmonized multimodal MRI acquisition; and (iii) integration with comprehensive multimodal clinical phenotyping and biobanking.

Mechanistically, BBB alterations in SSD are often linked to inflammatory and vascular pathways. Meta-analytic evidence indicates elevated peripheral pro-inflammatory markers, including IL-6, TNF-α, and IL-1β, across illness stages [19, 47, 48]. Furthermore, experimental work supports the link between inflammatory signaling and dysfunction in endothelial tight junctions and neurovascular unit, which may increase barrier permeability [5].

Within this framework, IMPACT is positioned to evaluate hypothesis-driven associations—particularly whether systemic inflammation relates to BBB leakage in a subgroup and whether BBB permeability metrics are associated with symptom dimensions, cognition, and longer-term functional trajectories.

In summary, IMPACT provides a structured longitudinal methodology to quantify BBB permeability in SSD using DCE-MRI and to integrate BBB measures with deep clinical and biological phenotyping. The study is designed to clarify the temporal profile and clinical and biological relevance of BBB dysfunction in SSD and to generate a data foundation for subsequent biomarker-informed stratification and mechanism-oriented hypotheses in precision psychiatry.

**Table 1.** Overview of baseline and follow-up examinations. The basic phenotyping of the Munich Mental Health Biobank has been described in detail elsewhere [24]. Abbreviations: ASL, Arterial Spin Labeling; BACS, Brief Assessment of Cognitive Function in Schizophrenia; CDSS, Calgary Depression Scale for Schizophrenia; CGI, Clinical Global Impression; DCE, Dynamic Contrast-Enhanced; GAF, Global Assessment of Functioning; PANNS, Positive and Negative Syndrome Scale; sMRI, Structural MRI; TMT, Trail Making Test.

|  | Study visit |  |  |
| --- | --- | --- | --- |
|  | Day 0<br>(baseline) | Day 28 - 14<br>days | V1 + 2.5 years<br>(+/- 90 days) |
| Assessments | V1 | V2 | V3 |
| **Clinical characterization** |  |  |  |
| Basic phenotyping Munich Mental Health Biobank and German Center for Mental Health (DZPG) | X |  |  |
| M.I.N.I. interview (DSM-5-TR) | X |  |  |
| Check inclusion/exclusion criteria | X | X | X |
| Psychiatric and medical history | X | X | X |
| Previous and current use of alcohol and illicit drugs | X | X | X |
| Current and previous medication | X | X | X |
| PANNS | X | X | X |
| PANNS RSWG criteria | X | X | X |
| CDSS | X |  |  |
| CGI-S | X | X | X |
| GAF | X | X | X |
| **Biomaterials** |  |  |  |
| Biobank blood sampling | X |  |  |
| Cerebrospinal fluid (optional) | X |  |  |
| Serum | X | X | X |

**Table 1. Overview of baseline and follow-up examinations.**
| Neuroanatomy |  |  |  |
| --- | --- | --- | --- |
| sMRI, rsMRI, ASL | X | X | X |
| DCE-MRI | X | X | X |

## Data Availability

All data produced in the present study will be made available upon reasonable request to the authors

## Funding

The procurement of the Prisma 3T MRI scanner was supported by the Deutsche Forschungsgemeinschaft (DFG, INST 86/1739-1 FUGG). VY was supported by the Residency/PhD track of the International Max Planck Research School for Translational Psychiatry (IMPRS-TP) and by the Faculty of Medicine at LMU Munich (FöFoLe Reg.-Nr. 1226/2024). JM was supported by the Faculty of Medicine at LMU Munich (FöFoLe Reg.-Nr. 1167) and by the Medical & Clinician Scientist Program (MCSP of the Faculty of Medicine, LMU Munich, Munich, Germany. No funding was received by commercial or not-for-profit sectors.

## Contributions

JM, DK and EW designed and conceptualized the Study. EW and JM wrote the protocol. VY, JM, EB, AW, AZ, IL, DL, CL, MS, and MB recruited patients and collected study data. LR, DK, and BS provided supervision. AW, AZ and JM wrote the first draft of the manuscript, and all authors commented on previous versions of the manuscript. All authors read and approved the final manuscript.

## Notes

### Competing Interest Statement

The authors have declared no competing interest.

### Author Declarations

Ethics committee/IRB of LMU University Hospital Munich gave ethical approval for this work

